# Towards mitigating health inequity via machine learning: a nationwide cohort study to develop and validate ethnicity-specific models for prediction of cardiovascular disease risk in COVID-19 patients

**DOI:** 10.1101/2023.09.13.23295489

**Authors:** Freya Allery, Marta Pineda-Moncusí, Christopher Tomlinson, Nikolas Pontikos, Johan H Thygesen, Sara Khalid, the CVD-COVID-UK/COVID-IMPACT Consortium

## Abstract

**Background:** Emerging data-driven technologies in healthcare, such as risk prediction models, hold great promise but also pose challenges regarding potential bias and exacerbation of existing health inequalities, which have been observed across diseases such as cardiovascular disease (CVD) and COVID-19. This study addresses the impact of ethnicity in risk prediction modelling for cardiovascular events following SARS-CoV-2 infection and explores the potential of ethnicity-specific models to mitigate disparities.

**Methods:** This retrospective cohort study utilises six linked datasets accessed through National Health Service (NHS) England’s Secure Data Environment (SDE) service for England, via the BHF Data Science Centre’s CVD-COVID-UK/COVID-IMPACT Consortium. Inclusion criteria were established, and demographic information, risk factors, and ethnicity categories were defined. Four feature selection methods (LASSO, Random Forest, XGBoost, QRISK) were employed and ethnicity-specific prediction models were trained and tested using logistic regression. Discrimination (AUROC) and calibration performance were assessed for different populations and ethnicity groups.

**Findings:** Several differences were observed in the models trained on the whole study cohort vs ethnicity-specific groups. At the feature selection stage, ethnicity-specific models yielded different selected features. AUROC discrimination measures showed consistent performance across most ethnicity groups, with QRISK-based models performing relatively poorly. Calibration performance exhibited variation across ethnicity groups and age categories. Ethnicity-specific models demonstrated the potential to enhance calibration performance for certain ethnic groups.

**Interpretation:** This research highlights the importance of considering ethnicity in risk prediction modelling to ensure equitable healthcare outcomes. Differences in selected features and asymmetric calibration across ethnicities underscore the necessity of tailored approaches. Ethnicity-specific models offer a pathway to addressing disparities and improving model performance. The study emphasises the role of data-driven technologies in either alleviating or exacerbating existing health inequalities.

**Evidence before this study:** Research has suggested that SARS-CoV-2 infections may have prognostic value in predicting later cardiovascular disease outcomes, two diseases where ethnicity-based health inequalities have been observed. Existing health inequalities are at risk of being exacerbated by bias in emerging data-driven technologies such as risk prediction models, and there currently exists no recommended practice to mitigate this issue. Model performances are not typically stratified by ethnic groups and, if reported, ethnic groups are often only included in higher-level categories that have been criticised for simplicity of definition and for missing key ethnic heterogeneity.

**Added value of this study:** This study demonstrates the impact of including an in-depth consideration of ethnicity and its granularity in risk prediction modelling for cardiovascular event prediction in patients following a SARS-CoV-2 infection. This is one of, if not the first, set of models specifically built for and representative of all ethnic groups across an entire population, evaluating different practices to best mitigate ethnicity-based disparities in prediction algorithms. Moreover, ethnicity data has historically not been well captured, with as many as 1 in 3 individuals missing ethnicity data in their health records. With data linkage, this work is the first to analyse 96% complete ethnicity records in one of the world’s largest ethnically diverse routinely collected datasets.

**Implications of all of the available evidence:** This study highlights the potential of tailoring feature selection, performance measures, and probability scores to different ethnic groups through ethnicity-specific risk prediction models to mitigate prediction bias. We identify differences between models trained on the global study populations to cohorts of specific ethnicities, and encourage the use of more granular ethnicity categories to capture the diversity of underlying populations. Such approaches will allow for newly developed data-driven tools to cater to the ethnic heterogeneity present between populations and ensure that emerging technologies translate into equitable health outcomes for everyone.

## Introduction

Ethnicity is a construct defined by multiple dimensions [1], from biological attributes such as skin colour to other descriptors such as country of birth, language, and religion [2]. An individual’s self-identified ethnic identity can have tangible implications across various sectors such as education, employment, and health [3][4][5]. Within the context of healthcare, disparities may be seen through access to care, disease prevalence and life expectancy [6]. The unequal impact of the COVID-19 pandemic on ethnic minority populations has furthered awareness of ethnicity-based health disparities [7][8][9], shown through higher rates of infection, severe illness, and death, with the death rate for people of Black African descent reported to be 3·5 times higher than for White people in the UK [10][11][12]. Similar disparities are found in cardiovascular disease (CVD), where mortality rates in South Asian communities were found to be 1.4 times higher than the general population in 2014 [13][14]. Recently, it has been identified that COVID-19 outcomes may have prognostic value in predicting patients at high risk of CVD, due to post-acute cardiovascular manifestations of COVID-19 [15].

To derive ethnicity-based insights in healthcare, electronic health records (EHRs) provide a unique opportunity to analyse ethnicity with a plethora of information, from prescribed medications and surgical interventions to other demographic information such as age and sex [16][17]. In England, ethnicity concepts in EHRs are most often stored either as one of 19 ethnicity codes available through Primary Code recordings informed by the Office of National Statistics (ONS) [18], or as 400+ codes defined by Systematized Nomenclature of Medicine - Clinical Terms (SNOMED-CT) from primary care providers [19]. However, the available granularity of ethnicity data is often underutilised in research, where data are typically collapsed into six categories (White, Black or Black British, Asian or Asian British, Mixed, Other Ethnic Group and Unknown) [20][21]. These six higher-level category groupings have been criticised as too broad and unspecific to capture important heterogeneity [20][22][23], which can have a real impact on representation in research [24].

The nature of our healthcare systems is undergoing a significant paradigm shift, with the increasing capacities of technology facilitating data-driven innovation based on complex, multi-modal patient data [25][26]. There is currently a huge amount of traction behind the use of machine learning algorithms to predict the risk of patient outcomes based on a set of salient, patient-specific features which are commonly derived from routinely collected EHR data [27]. However, if bias is present in the data or in the algorithms then there is a risk of incorrect care, or no care, being given to a patient and several clinically deployed risk prediction algorithms have been found to have ethnicity-based biases [28][29]. This is emblematic of the potential dangers that data-driven developments pose to health equity and highlight the importance of considering ethnic heterogeneity in their development.

Through the lens of incident CVD prediction in COVID-19 patients, this study aims to examine the differences between risk prediction models where feature selection and model training has been carried out on the general study population compared to ethnicity-specific models where feature selection and model training has been carried out separately in specific ethnic groups.

## Methods

### Study design and data sources

In this retrospective cohort study, data was extracted from the National Health Service (NHS) England and accessed through NHS England’s Secure Data Environment (SDE) service for England, via the BHF Data Science Centre’s CVD-COVID-UK/COVID-IMPACT Consortium where individuals have been previously pseudonymised and linked across the different datasets included in the environment.

Our predictive models were built using data from six of the linked datasets, including the national laboratory COVID-19 testing data from Public Health England, Second Generation Surveillance System (SGSS); primary care data from the General Practice Extraction Service Extract for Pandemic Planning and Research (GDPPR); hospital admission data from Secondary Uses Service (SUS); Hospital Episode Statistics for admitted patient care (HES-APC); COVID-19 hospital admission data from the COVID-19 Hospitalisations in England Surveillance System (CHESS; a dataset curated at the start of the pandemic to provide information about COVID-19 hospitalisations); and mortality information from the Office for National Statistics (ONS) Civil Registration of Deaths [30] [31]. This included patient demographics, disease phenotype diagnosis, medication, positive SARS-CoV-2 test, COVID-19 hospitalisation and COVID-19 death.Our predictive models were built using data from six of the linked datasets, including the national laboratory COVID-19 testing data from Public Health England, Second Generation Surveillance System (SGSS); primary care data from the General Practice Extraction Service Extract for Pandemic Planning and Research (GDPPR); hospital admission data from Secondary Uses Service (SUS); Hospital Episode Statistics for admitted patient care (HES-APC); COVID-19 hospital admission data from the COVID-19 Hospitalisations in England Surveillance System (CHESS; a dataset curated at the start of the pandemic to provide information about COVID-19 hospitalisations); and mortality information from the Office for National Statistics (ONS) Civil Registration of Deaths [30] [31]. This included patient demographics, disease phenotype diagnosis, medication, positive SARS-CoV-2 test, COVID-19 hospitalisation and COVID-19 death.

**Figure 1:**
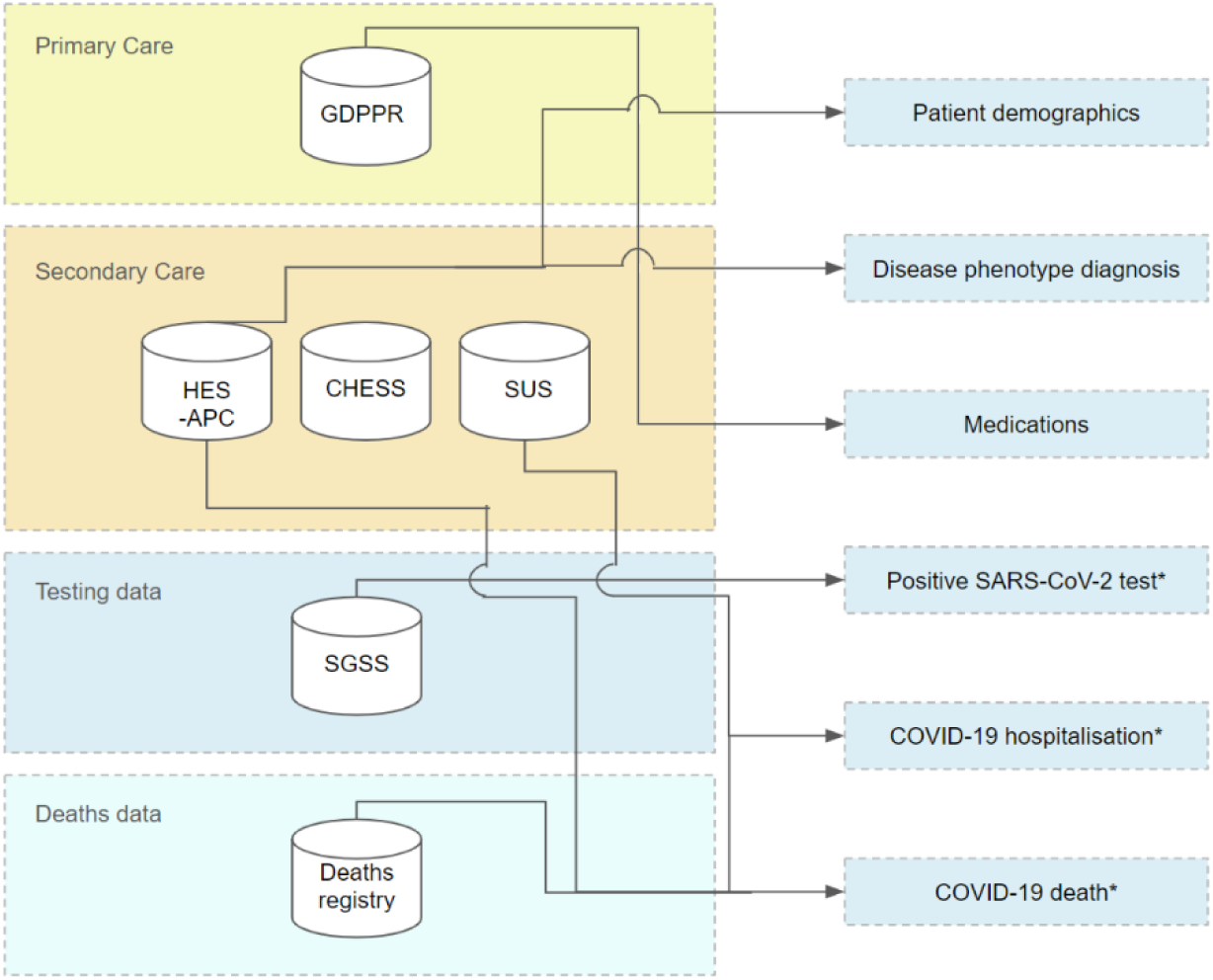
Data sources from NHS England’s SDE and the variables they inform. Patient demographics and disease phenotype diagnoses were derived from GDPPR and HES-APC, medications were derived from GDPPR, a positive SARS-CoV-2 test was derived from GDPPR and SGSS, COVID-19 hospitalisations were derived from CHESS and SUS data and COVID-19 death was derived from Deaths Registry data. *Indicates that variables were derived using definitions from Thygesen et al 2022 [31].

### Study population

All participants with a unique, de-identified patient ID were considered against the following inclusion criteria: a) at least one record in GDPPR, b) first positive COVID-19 infection between the 30^th^ September 2020 and the 12^th^ February 2021 (index date: date of their confirmed COVID-19 infection), c) over the age of 18 at the index date and d) more than a year of records available from their index date. The period restriction for positive COVID-19 infections was chosen to avoid confounding effects of pandemic waves, variants and other time-varying factors.

Individuals were included from their index date and followed to the earliest end point: i) outcome of study, ii) end of study date or iii) death date. The study end date was June 29^th^, 2022.

### Outcomes

The primary outcome was a CVD event within 1 year of index date (CVD-1y). These events were identified by an and International Classification of Diseases, Tenth Revision (ICD-10) or SNOMED-CT record (in hospital data and GDPPR data, respectively) for a CVD event, including diagnoses of coronary heart disease events, stroke and transient ischaemic attack. Codelists were derived from Kuan et al. 2019 [32]. As a secondary outcome, we also explored CVD events within 30 days, 90 days, 180 days and 2 years of the index date.

A secondary outcome was COVID-19 death, defined as either (i) record of ICD-10 term for suspected or confirmed COVID-19 diagnosis on the death certificate, (ii) death within 28 days from first recorded COVID-19 event (positive test, diagnosis, or admission), or (iii) hospital admission due to COVID-19 with a discharge method or destination indicating death, regardless of cause and duration after the index event.

### Demographic variables and risk factors

The participant’s demographic information included age, sex, ethnicity, and socio-economic deprivation (patient-level index of multiple deprivation (IMD) quintile where 1 indicates most deprived and 5 indicates the least deprived). Age and sex were derived from the most recent non-missing value across primary care (GDPPR) and secondary care (HES-APC). For ethnicity, prioritisation was given to the most recent primary care record, with secondary care records used where data was missing or coded as unknown.

The analysed risk factors were informed by the QRISK3 study alongside other commonly used risk factors for COVID-19 severity and CVD [33]. Risk factors included: obesity, history of CVD, CVD 1 year prior to index date, smoking, cancer, alcohol problems, hypertension, alcoholic liver disease, atrial fibrillation (AF), chronic kidney disease (CKD), rheumatoid arthritis (RA), schizophrenia, bipolar disorder, severe depression, chronic obstructive pulmonary disease (COPD), erectile dysfunction, dementia, diabetes, prior fractures (hip and wrist), osteoporosis, dementia, diabetes, statins, anti-hypertensive drugs, anti-psychotic medication, anti-coagulant drugs, anti-diabetic drugs and anti-platelet drugs. A severe mental illness variable was created by combining schizophrenia, bipolar disorder and severe depression features from the curated dataset.

### Ethnicity categories

Ethnicity was recorded in the EHRs through nineteen NHS Primary Code ethnicity groups (British, Irish, Any other White background, White and Black Caribbean, White and Black African, White and Asian, Any other Mixed background, Indian, Pakistani, Bangladeshi, Any other Asian background, Caribbean, African, Any other Black background, Chinese, Any other ethnic group, Gypsy or Irish Traveller, Arab, Unknown or not stated), which are defined by the NHS England Data Dictionary. These ethnicity concepts were also mapped to one of six higher-level groupings (White, Black or Black British, Asian or Asian British, Mixed, Other ethnic group and Unknown or not stated).

### Statistical analysis

Characteristics of individuals included in the study were described at index date stratified by their Primary Code ethnicity groups.

#### Feature selection

Four methods were used for feature selection:1) least absolute shrinkage and selection operator (LASSO) Regression, 2) Random Forest Classifier, 3) Extreme Gradient Boosting (XGBoost) Classifier, and as a reference method 4) the list of features defined in the QRISK3 algorithm, available in the dataset [33]. Hyperparameter tuning included repeated stratified 10-fold cross-validation with 10 repeats.

#### Prediction modelling

Prediction models were individually trained using different sub-groups of the cohort. Each set of prediction models comprised of four Logistic Regression models, differentiated by the use of one of the four feature selection methods. Each set of models were trained on the following population groups: a) the whole general study population, b) ethnicity-specific populations for each of the six higher-level ethnicity categories and c) ethnicity-specific populations separated by the nineteen Primary Code ethnicity categories. A random 80:20 train/test split was used for all population groups, and a visualisation of the process is shown in Figure 2.

**Figure 2:**
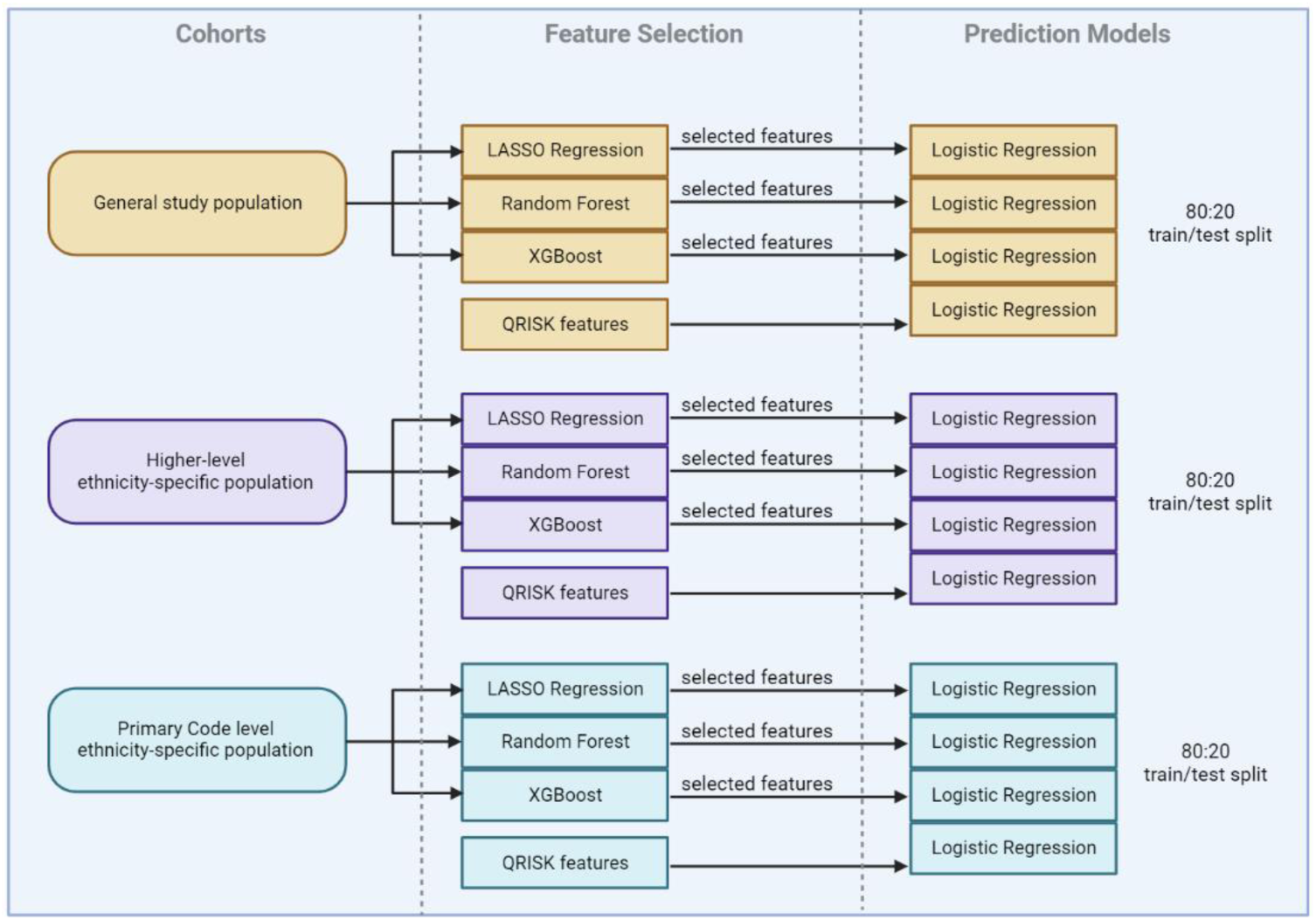
Modelling process indicating the different cohorts the models were trained on, the feature selection stage and the implemented prediction models. The cohorts the models were trained and tested on include the general study population, the 6 Higher-level ethnicity-specific populations and the 19 Primary Code populations. For each of these populations, feature selection was carried out using LASSO Regression, Random Forest, XGBoost. Additionally, features as defined in QRISK were also used, and each set of features were fed into 4 separate Logistic Regression Models to carry out the prediction task. All processes were carried out with a random 80:20 train/test split.

#### Performance assessment

In the testing sets, the discrimination of the Logistic Regression models was assessed using the area under the receiver operator characteristic (AUROC) and the calibration was assessed via a calibration curve.

Data cleaning, pre-processing, phenotype identification, and cohort creation were performed using Python (version 3.7) and Spark SQL (version 2.4.5) on Databricks (version 6.4). Prediction modelling and analysis were conducted using Python (version 3.7) using skit-learn (version 1.0). Summary statistics were created in RStudio (Professional version 1.3.1093.1 driven by R Version 4.0.3) using tableOne (version 0.12.0). Figures were constructed using the ggplot2 (version 3.3.3) package and plotly (version 4.10.2).

This analysis was performed according to a pre-specified analysis plan published on GitHub, along with the phenotyping and analysis code (https://github.com/BHFDSC/CCU037_03).

### Role of the funding source

The funders had no role in study design, data collection, data analysis, data interpretation of data, or writing of the report.

## Results

### Study population

There were 62,345,357 unique patient IDs present in the GDPPR dataset up until the 29th of June 2022. 59,475,405 individuals were excluded due to an absence of positive test within the defined wave 2 dates, 110,098 were excluded for having less than 1 year of records available prior to index date and 355,910 were excluded whose age was <18 at the index date. A further 167 individuals were excluded on account of having a date of death before their index date, as this indicated data quality issues. The final study population consisted of 2,423,777 who met the study’s inclusion criteria.

### Feature selection

Out of the employed feature selection methods and the 51 possible features available, LASSO Regression was the least selective, followed by Random Forest and XGBoost, which selected the fewest features. For the methods trained on the global cohort, LASSO selected 44 features, Random Forest selected 16 features and XGBoost selected 6 features. When trained on Higher-level ethnicity groups the average number of features selected were 29.8 ± 7.0 by LASSO, 19.0 ± 2.0 by Random Forest and 5.8 ± 1.2 by XGBoost. For LASSO, Random Forest and XGBoost models when trained on Primary Code ethnicity groups, the average number of features selected were 22.4 ± 9.0, 16.3 ± 2.3 and 7.5 ± 4.2, respectively. A visualisation of the different features selected by XGBoost for each of the groups is given in Figure 3. The model trained on the global cohort selected the age group features 18-29, 30-39, 40-49 and 70-79, CVD history and CVD history one year prior to COVID-19 infection. When trained on specific ethnicity groups, other features were selected such as statins in the Bangladeshi population, hypertension in the Caribbean population and diabetes in the Chinese population.

**Figure 3:**
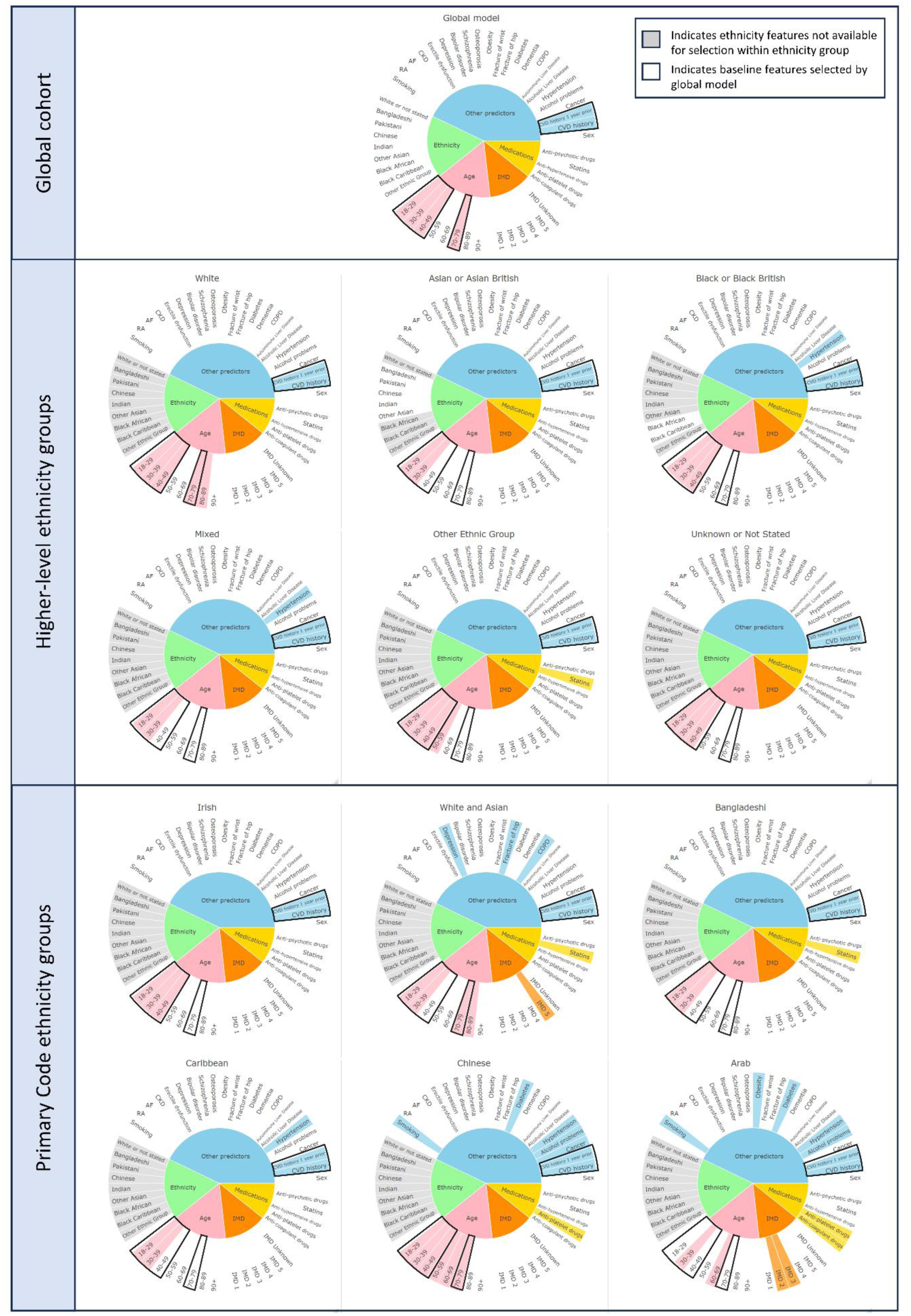
Features selected by XGBoost, as stratified by Primary Code ethnicity group. Shown are chart segments showing age, index of multiple deprivation (IMD), ethnicity, medications and other predictors. The coloured segments represent the features that have been selected by the methods and the blank segments indicate that the feature was not selected for inclusion in prediction.

### Discrimination

The AUROC discrimination measures were calculated for each of the four models for the held-out testing data and each ethnicity grouping. The models were Logistic Regression using features selected by LASSO (LASSO + LR), QRISK (QRISK + LR), Random Forest (RF + LR) and XGBoost (XGB + LR) and the population stratifications were the global cohort, the 6 Higher-level ethnicity groupings and the 19 Primary Code ethnicity groupings. For the model trained on the global cohort, the highest AUROC across the Primary Code ethnicity groups was for LASSO + LR in the White and Asian cohort 0.952 [0.917 – 0.988] and the lowest was 0.778 [0.636 – 0.920] for RF+ LR in the Arab cohort. In all ethnicity groups except for Arab, QRISK + LR was the poorest performing model. For the models trained on specific higher-level ethnicity groups, the highest performance was 0.942 [0.910 – 0.975] for LASSO + LR in the White and Black Caribbean group and the lowest was 0.783 [0.586 – 0.981] for RF + LR in the Gypsy and Irish Traveller population. QRISK + LR was the lowest performing across all groups except for Gypsy or Irish Traveller, White and Asian, Chinese and Unknown populations. For the models trained on the Primary Code ethnicity groups, the highest performance was 0.957 [0.919 – 0.995] in LASSO + LR for the Gypsy and Irish Traveller population and the lowest performance was 0.775 [0.636 – 0.913] in QRISK + LR for the Arab population. QRISK + LR was the lowest performing in all groups except for the White and Black Caribbean and the Unknown populations. Figure 4 shows a visualisation for the AUROC for each set of models, stratified by Primary Code ethnicity group.

**Figure 4:**
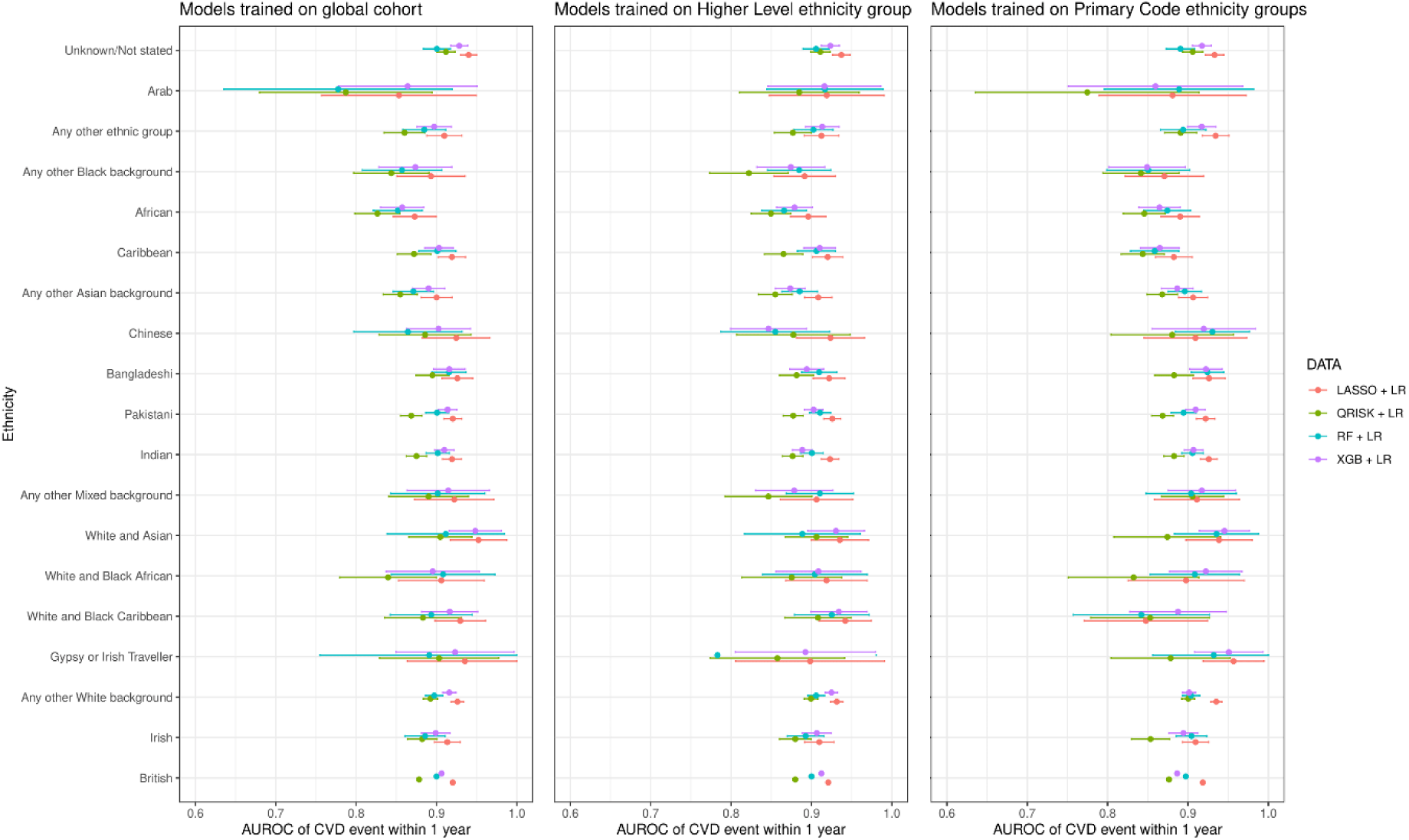
AUROC for the models trained on the global cohort, Higher Level ethnicity groups and Primary Code ethnicity groups, stratified by Primary Code ethnicity group. The models shown are Logistic Regression using features selected by LASSO (LASSO + LR), QRISK (QRISK + LR), Random Forest (RF + LR) and XGBoost (XGB + LR), indicated by red, green, blue, and purple, respectively. Shown are the AUROC values with bars representing the 95% confidence intervals.

### Calibration

For the calibration measures, the models trained on the global cohort showed inconsistent performance between ethnicities, performing best for White and Unknown ethnic groups, and significantly poorer for the ethnic minority groups, especially in older age groups where there were wide confidence intervals on the estimates. Good calibration performance can be observed in British, Irish, Any other White background, Indian, Caribbean and Unknown populations, whilst the remaining ethnicity groups exhibit poorer performances, particularly for age > 70. Several models trained on ethnicity-specific sub-cohorts were implemented to evaluate any change in performances and this resulted in overall improvements in model calibration, particularly when deploying ethnicity-specific models at a Primary Code ethnicity category level. A visualisation for the model calibration when trained on Primary Code ethnicity groups is presented in Figure 5.

**Figure 5:**
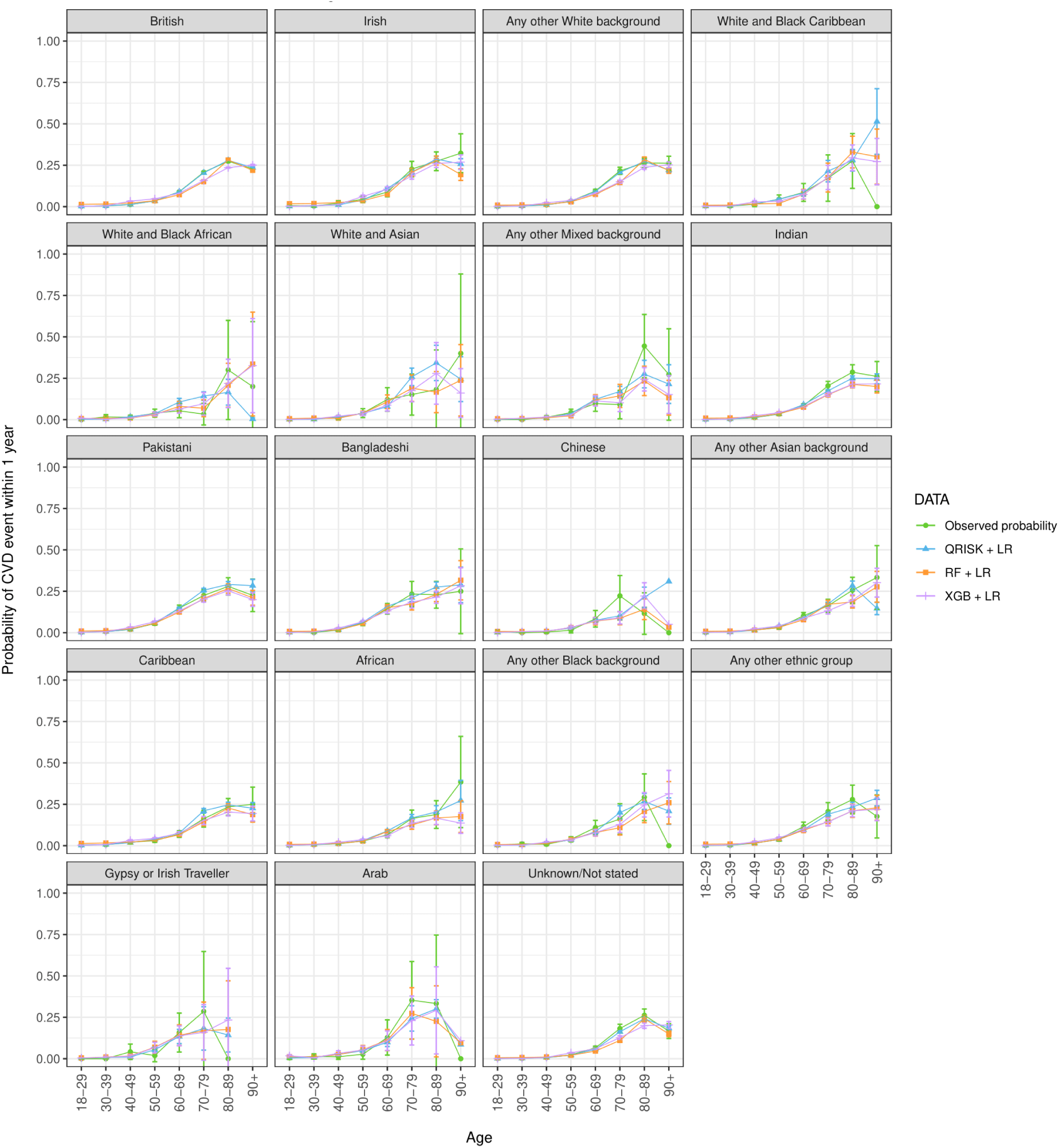
AUROC for the models trained on Primary Code ethnicity groups, stratified by Primary Code ethnicity group and age deciles. Shown is the observed probability in green and the predicted probabilities from the Logistic regression models using features selected QRISK (QRISK + LR), Random Forest (RF + LR) and XGBoost (XGB + LR), indicated by blue, orange, and purple lines, respectively. The bars indicate the 95% confidence intervals.

## Discussion

This study explored the usage of low-level specific ethnicity data than typically used in EHR studies to evaluate the impact this may have on performance of risk prediction models on different ethnicities through the lens of post COVID-19 infection CVD risk prediction on a population scale. Further to this, ethnicity-specific models were implemented to explore their potential in mitigating observed disparities.

The differences in selected features for models trained on ethnicity-specific groups vs models trained on the overall sample exemplifies the differences of health needs and outcomes between ethnicities and shows the importance of incorporating this consideration at the developmental stage of machine learning algorithms. Moreover, although the AUROC discrimination measures show little variation, the asymmetric calibration performance between ethnicity groups highlights a need to further consider the efficacy of data-driven prediction tasks across all populations. The introduction of models trained on ethnicity-specific groups demonstrated their potential to improve performance for some ethnicity groups, for example in Pakistani, Bangladeshi and Arab populations. These insights give motivation for the impact of research on under-represented groups to be considered at the forefront of future studies, particularly in health outcomes known to contain existing inequities.

This study is, to the best of our knowledge, the most in-depth exploration of ethnicity-stratified performances in prediction modelling. The implementation of ethnicity-specific risk prediction tools is a potential avenue to achieve comparable performances across ethnicities but requires further analysis into potential issues introduced by such models. Future work should include a more in-depth exploration into the very granular SNOMED-CT ethnicity codes and their potential in research, as well as further investigation into the mechanisms underpinning poorer model performance in ethnic minorities and recommendations to mitigate such effects. Moreover, validating these models on an external data set would enable the ethnicity-based performances of the models to be concluded within a separate population and give further confidence in these results.

A better understanding is required of the nature and use of key demographic indicators such as ethnicity data within EHRs to ensure that emerging research works to alleviate, rather than perpetuate, inequalities that are embedded in healthcare structures. With the steep rise in data-driven health research, the promise of such technologies in providing personalised, patient-specific care is only attainable if it is ensured that insights are representative of all individuals in the target population. It is paramount that poorer performances for already marginalised groups in the healthcare system, such as ethnic minorities, are not overlooked; or there is a real risk of data-driven innovation exacerbating existing health inequalities.

## Conclusion

In conclusion, our study underscores the pivotal role of ethnicity in shaping risk prediction modelling for cardiovascular events post-SARS-CoV-2 infection. As data-driven healthcare technologies advance, so too does the urgency to address inherent biases and disparities. Enabled by six linked NHS England datasets, we illuminate the intricate relationship between ethnicity, predictive features, and model efficacy. Notably, our investigation into ethnicity-specific models presents a promising avenue for narrowing performance gaps. By refining models to cater to specific ethnic groups, we glimpse a path toward more equitable predictive accuracy. This research resonates as a clarion call for embracing inclusivity and equity in healthcare prediction, safeguarding that technological advancements translate to improved health outcomes for all, regardless of their ethnic background.

## Funding

The British Heart Foundation Data Science Centre (grant No SP/19/3/34678, awarded to Health Data Research (HDR) UK) funded co-development (with NHS England) of the Secure Data Environment service for England, provision of linked datasets, data access, user software licences, computational usage, and data management and wrangling support, with additional contributions from the HDR UK Data and Connectivity component of the UK Government Chief Scientific Adviser’s National Core Studies programme to coordinate national COVID-19 priority research. Consortium partner organisations funded the time of contributing data analysts, biostatisticians, epidemiologists, and clinicians.

## Acknowledgments

This work is carried out with the support of the BHF Data Science Centre led by HDR UK (BHF Grant no. SP/19/3/34678). This study makes use of de-identified data held in NHS England’s Secure Data Environment service for England and made available via the BHF Data Science Centre’s CVD-COVID-UK/COVID-IMPACT consortium. This work uses data provided by patients and collected by the NHS as part of their care and support. We would also like to acknowledge all data providers who make health relevant data available for research.

## Authors’ roles

Conceptualisation: SK, DPA, AD, GC. Data curation: FA, MPM, CT, JT. Formal analysis: FA, MPM, TB. Funding acquisition: SK. Data interpretation: FA, MPM, JT, SK. Writing original draft: FA, MPM, JT, SK. Writing review and editing: all authors. Approving final version of manuscript: all authors. SK, FA and MPM take responsibility for the integrity of the data analysis. CS is the Director of the BHF Data Science Centre and coordinated approvals for and access to data within NHS England’s Secure Data Environment service for England for CVD-COVID-UK/COVID-IMPACT.

## Conflicts of interest

No conflicts of interest.

## Ethical approval

The North East - Newcastle and North Tyneside 2 research ethics committee provided ethical approval for the CVD-COVID-UK/COVID-IMPACT research programme (REC no: 20/NE/0161) to access, within secure trusted research environments, unconsented, whole-population, de-identified data from electronic health records collected as part of patients’ routine healthcare.

Our project (proposal CCU037, short title: *Minimising bias in ethnicity data*) agreed the objectives of the consortium’s ethical and regulatory approvals, and was authorized by the BHF Data Science Centre’s Approvals and Oversight Board. Approved researchers (MPM, AD, SK) conducted the analyses within the NHS England’s SDE via secure remote access. Ensuring the anonymity of individuals, only summarized-aggregated results that were manually reviewed by the NHS England ‘safe outputs’ escrow service were exported from the SDE.

## Data sharing

The data used in this study are available in NHS England’s Secure Data Environment (SDE) service for England, but as restrictions apply they are not publicly available (https://digital.nhs.uk/services/secure-data-environment-service). The CVD-COVID-UK/COVID-IMPACT programme, led by the BHF Data Science Centre (https://bhfdatasciencecentre.org/), received approval to access data in NHS England’s SDE service for England from the Independent Group Advising on the Release of Data (IGARD) (https://digital.nhs.uk/about-nhs-digital/corporate-information-and-documents/independent-group-advising-on-the-release-of-data) via an application made in the Data Access Request Service (DARS) Online system (ref. DARS-NIC-381078-Y9C5K) (https://digital.nhs.uk/services/data-access-request-service-dars/dars-products-and-services). The CVD-COVID-UK/COVID-IMPACT Approvals & Oversight Board (https://bhfdatasciencecentre.org/areas/cvd-covid-uk-covid-impact/) subsequently granted approval to this project to access the data within NHS England’s SDE service for England. The de-identified data used in this study were made available to accredited researchers only. Those wishing to gain access to the data should contact in the first instance.

## Notes

### Competing Interest Statement

The authors have declared no competing interest.

### Author Declarations

The North East - Newcastle and North Tyneside 2 research ethics committee provided ethical approval for the CVD-COVID-UK/COVID-IMPACT research programme (REC no: 20/NE/0161) to access, within secure trusted research environments, unconsented, whole-population, de-identified data from electronic health records collected as part of patients' routine healthcare. Our project (proposal CCU037, short title: Minimising bias in ethnicity data) agreed the objectives of the consortium's ethical and regulatory approvals, and was authorized by the BHF Data Science Centre's Approvals and Oversight Board. Approved researchers (MPM, AD, SK) conducted the analyses within the NHS England's SDE via secure remote access. Ensuring the anonymity of individuals, only summarized-aggregated results that were manually reviewed by the NHS England safe outputs escrow service were exported from the SDE.

### Summary of Updates

Added additional authors to the author list. No other changes have been made.

## References

[1] S. Dein, “Race, Culture and Ethnicity in Minority Research: A Critical Discussion,” J Cult Divers, vol. 13, no. 2, pp. 68–75, 2006.

[2] J. Burton, A. Nandi, and L. Platt, “Measuring ethnicity: Challenges and opportunities for survey research,” Ethn Racial Stud, vol. 33, no. 8, pp. 1332–1349, 2010, doi: 10.1080/01419870903527801.

[3] J. T. E. Richardson, “The attainment of ethnic minority students in UK higher education,” Studies in Higher Education, vol. 33, no. 1, pp. 33–48, Feb. 2008, doi: 10.1080/03075070701794783.

[4] G. V. F. Miller and C. J. Travers, “Ethnicity and the experience of work: Job stress and satisfaction of minority ethnic teachers in the UK,” International Review of Psychiatry, vol. 17, no. 5, pp. 317–327, Oct. 2005, doi: 10.1080/09540260500238470.

[5] R. A. Hackett, A. Ronaldson, K. Bhui, A. Steptoe, and S. E. Jackson, “Racial discrimination and health: a prospective study of ethnic minorities in the United Kingdom,” BMC Public Health, vol. 20, no. 1, Dec. 2020, doi: 10.1186/s12889-020-09792-1.

[6] E. Wilkinson and G. Randhawa, “An examination of concordance and cultural competency in the diabetes care pathway: South Asians living in the United Kingdom,” Indian J Nephrol, vol. 22, no. 6, pp. 424–430, 2012, doi: 10.4103/0971-4065.106033.

[7] D. Cucinotta and M. Vanelli, “WHO declares COVID-19 a pandemic,” Acta Biomedica, vol. 91, no. 1, pp. 157–160, 2020, doi: 10.23750/abm.v91i1.9397.

[8] D. Pan et al., “The impact of ethnicity on clinical outcomes in COVID-19: A systematic review,” EClinicalMedicine, vol. 23, Jun. 2020, doi: 10.1016/j.eclinm.2020.100404.

[9] D. B. G. Tai, A. Shah, C. A. Doubeni, I. G. Sia, and M. L. Wieland, “The Disproportionate Impact of COVID-19 on Racial and Ethnic Minorities in the United States,” Clinical Infectious Diseases, vol. 72, no. 4, pp. 703–706, 2021, doi: 10.1093/cid/ciaa815.

[10] T. S. Gaynor and M. E. Wilson, “Social Vulnerability and Equity: The Disproportionate Impact of COVID-19,” Public Adm Rev, vol. 80, no. 5, pp. 832–838, 2020, doi: 10.1111/puar.13264.

[11] T. Kirby, “Evidence mounts on the disproportionate effect of COVID-19 on ethnic minorities,” Lancet Respir Med, vol. 8, no. 6, pp. 547–548, Jun. 2020, doi: 10.1016/S2213-2600(20)30228-9.

[12] V. Abedi et al., “Racial, Economic, and Health Inequality and COVID-19 Infection in the United States,” J Racial Ethn Health Disparities, vol. 8, pp. 732–742, 2020, doi: 10.1007/s40615-020-00833-4.

[13] Z. Jalal, · Sotiris Antoniou, · David Taylor, V. Paudyal, K. Finlay, and · Felicity Smith, “South Asians living in the UK and adherence to coronary heart disease medication: a mixed-method study,” Int J Clin Pharm, vol. 41, pp. 122–130, 2014, doi: 10.1007/s11096-018-0760-3.

[14] S. Bellary et al., “Premature cardiovascular events and mortality in south Asians with type 2 diabetes in the United Kingdom Asian Diabetes Study effect of ethnicity on risk,” Curr Med Res Opin, vol. 26, no. 8, pp. 1873–1879, 2010, doi: 10.1185/03007995.2010.490468.

[15] Y. Xie, E. Xu, B. Bowe, and Z. Al-Aly, “Long-term cardiovascular outcomes of COVID-19,” Nat Med, 2022, doi: 10.1038/s41591-022-01689-3.

[16] J. Russel, “Electronic health records,” Chartered Society of Physiotherapy, 2020. https://www.csp.org.uk/professional-clinical/digital-physiotherapy/electronic-health-records (accessed Sep. 30, 2022).

[17] R. E. Gliklich, M. B. Leavy, and N. A. Dreyer, “Tools and Technologies for Registry Interoperability, 2nd addendum of Registries for Evaluating Patient Outcomes, A User’s Guide,” Dec. 2019.

[18] “Hospital Episode Statistics Data Dictionary - NHS Digital,” NHS Digital, 2021. https://digital.nhs.uk/data-and-information/data-tools-and-services/data-services/hospital-episode-statistics/hospital-episode-statistics-data-dictionary (accessed Sep. 30, 2022).

[19] NHS Digital, “SNOMED CT,” 2021.

[20] K. Khunti, A. Routen, A. Banejeree, and M. Pareek, “The need for improved collection and coding of ethnicity in health research,” J Public Health (Bangkok), vol. 3, no. 3, pp. 270–272, 2020, doi: 10.1093/pubmed/fdaa198.

[21] P. Kumarapeli, R. Stepaniuk, S. de Lusignan, R. Williams, and G. Rowlands, “Ethnicity recording in general practice computer systems,” J Public Health (Bangkok), vol. 28, no. 3, pp. 283–287, 2006, doi: 10.1093/pubmed/fdl044.

[22] K. Sultana and A. Sheikh, “Most UK datasets of routinely collected health statistics fail to collect information on ethnicity and religion,” J R Soc Med, vol. 101, pp. 463–465, 2008, doi: 10.1258/jrsm.2008.080007.

[23] Z. Tippu et al., “Ethnicity recording in primary care computerised medical record systems: an ontological approach,” J Innov Health Inform, vol. 23, no. 4, 2017, doi: 10.14236/jhi.v23i4.920.

[24] K. E. Kaseniit, I. S. Haque, J. D. Goldberg, L. P. Shulman, and D. Muzzey, “Genetic ancestry analysis on 93,000 individuals undergoing expanded carrier screening reveals limitations of ethnicity-based medical guidelines,” Genetics in Medicine, vol. 22, no. 10, pp. 1694–1702, 2020, doi: 10.1038/s41436-020-0869-3.

[25] F. Jiang, Y. Jiang, H. Zhi, and Y. Dong, “Artificial intelligence in healthcare: past, present and future,” Stroke Vasc Neurol, vol. 17, no. 2, 2017, doi: 10.1136/svn-2017-000101.

[26] A. I. Khan and A. Al-Badi, “Emerging data sources in decision making and AI,” in *Procedia Computer Science*, Elsevier B.V., 2020, pp. 318–323. doi: 10.1016/j.procs.2020.10.042.

[27] V. Sharma, I. Ali, S. van der Veer, G. Martin, J. Ainsworth, and T. Augustine, “Adoption of clinical risk prediction tools is limited by a lack of integration with electronic health records,” BMJ Health Care Inform, vol. 28, 2021, doi: 10.1136/bmjhci-2020-100253.

[28] Z. Obermeyer, B. Powers, C. Vogeli, and S. Mullainathan, “Dissecting racial bias in an algorithm used to manage the health of populations,” Science (1979), vol. 366, pp. 447–453, 2019, doi: 10.1126/science.aax2342.

[29] J. Feagin and Z. Bennefield, “Systemic racism and U.S. health care,” Soc Sci Med, vol. 103, pp. 7–14, Feb. 2014, doi: 10.1016/j.socscimed.2013.09.006.

[30] A. Wood, R. Denholm, S. Hollings, J. Cooper, and S. Ip, “Linked electronic health records for research on a nationwide cohort of more than 54 million people in England: data resource on behalf of the CVD-COVID-UK consortium,” BMJ, vol. 6, 2021, doi: 10.1136/bmj.n826.

[31] J. Thygesen, C. Tomlinson, S. Hollings, M. A. Mizani, and A. Handy, “COVID-19 trajectories among 57 million adults in England: a cohort study using electronic health records,” The Lancet, pp. 542–557, 2022, doi: 10.1016/S2589-7500(22)00091-7.

[32] V. Kuan et al., “A chronological map of 308 physical and mental health conditions from 4 million individuals in the English National Health Service,” Lancet Digit Health, vol. 1, no. 2, 2019, doi: 10.1016/S2589-7500(19)30012-3.

[33] J. Hippisley-Cox, C. Coupland, and P. Brindle, “Development and validation of QRISK3 risk prediction algorithms to estimate future risk of cardiovascular disease: Prospective cohort study,” The BMJ, vol. 357, 2017, doi: 10.1136/bmj.j2099.

